# COVID-19 excess death rate in Eastern European countries associated with weaker regulation implementation and lower vaccination coverage

**DOI:** 10.1101/2022.02.06.22270549

**Authors:** Alban Ylli, Genc Burazeri, Yan Yan Wu, Tetine Sentell

**Affiliations:** Department of Public Health, Faculty of Medicine, University of Medicine, Tirana, Albania; Institute of Public Health, Tirana, Albania; Department of International Health, School CAPHRI (Care and Public Health Research Institute), Maastricht University, Maastricht, The Netherlands; Office of Public Health Studies, University of Hawai‘i at Mānoa, Honolulu, Hawaii, USA

**Author notes:** **Corresponding author:** Alban Ylli, MD, PhD; Address: Rr. Aleksander Moisiu, No. 80, Tirana, Albania.

**Keywords:** COVID-19, European Region, excess mortality, regulation enforcement, rule of law, vaccination coverage

## Abstract

**Aim:** The objective of this analysis was to assess the association of excess COVID-19 mortality with regulation enforcement and vaccination rate in selected countries.

**Methods:** This analysis included 50 countries pertinent to the WHO European Region, in addition to USA and Canada. Excess mortality and vaccination data were retrieved from *“Our World In Data”* database, while regulation implementation was measured from a well-respected, standardized measure. Outpatient visits were also included in the analysis. Multiple linear regression was used to assess the independent association between excess mortality and each covariate.

**Results:** Excess mortality increased by 4.1/100 000 for every percent decrease in vaccination rate and with 6/100 000 for every decreased unit in the regulatory implementation score a country achieved in the Rule of Law Index.

**Conclusion:** Degree of regulation enforcement, likely including public health measure enforcement, may be an important factor in controlling COVID-19’s deleterious health impacts.

## Introduction

COVID-19 incidence and confirmed death rate are the most widely used epidemiological parameters to describe differences between countries in the level of risk and health outcomes in the pandemic (1). Both outcomes might be highly affected by countries’ testing and data collection capacities (2). Excess death rate is a more complex indicator but it may show the true impact of the pandemic across a population, especially in the long term (3). It may also serve as a better comparative measure across countries with differences in health system and surveillance resources (4).

In the countries of Eastern Europe, where intensive circulation of COVID 19 started later than in the West, public health measures were very effective in first half of 2020. Excess death rates in most of these countries were negligent in a time when most of Western Europe exceeded expected rates by 15%-35% (5). During the second wave in autumn-winter 2020, excess mortality in Eastern Europe was higher than most Western Europe and the difference has continued throughout 2021 (6). The countries of Eastern Europe started vaccination campaigns in a similar time frame with Western Europe, but the vaccination rates there have been comparatively lower (7).

Comparing and quantifying cumulative COVID impacts across country-level public health metrics can be challenging from an outcomes and a public health policy perspective. Efforts to measure the association of COVID-19 incidence with vaccination rate (8) have results that imply lack of efficacy of vaccination programs. Excess mortality may provide a better indicator of measurement of long-term health impact differences across countries, especially in the context of different vaccination rates. Publications analysing differences in the effectiveness of government interventions exist (9), but they take for granted the full implementation of the measures in various countries. Differences in enforcement of measures between countries are a known challenge for international analysis of country response measures to COVID-19 (10,11).

Government ‘regulatory enforcement’ levels generally likely impact the degree of public health measures enforcement and can thus serve as a proxy for this measurement. ‘Regulatory enforcement’ is one components of the Rule of Law index, calculated every year by World of Justice Project (WJP). It is a standardised estimate about how well government regulations are implemented for situations such as environmental restrictions, public health requirements, workplace safety conditions, business activity etc (12). Regulatory enforcement may thus be a potential factor in explaining country differences in pandemic outcomes.

The objective of this analysis was to assess the association of excess COVID-19 mortality with vaccination rate and regulation enforcement, controlling for health care utilization. We hypothesized that higher excess mortality rates in Eastern European countries are associated with lower ‘regulatory enforcement’ scores, which impact both government non-pharmaceutical interventions and vaccination programs. Thus, higher excess mortality can be explained by poorer implementation of public health measures and vaccination programs.

## Methods

This analysis included 50 countries pertinent to WHO European Region, in addition to USA and Canada. Excess mortality was calculated as observed deaths minus expected deaths per 100 000 population. Expected deaths were estimated from the average 5-year observation of country reported deaths. Excess mortality is for the period from 1 January 2020 to 2 January 2022. Excess mortality data were retrieved from “Our World In Data” database (13).Vaccination rate was calculated as the percentage of the total population who have received at least two doses of vaccine as reported by national programmes. The data are for the period 10-15 January 2022 (14).

The regulation implementation indicator was based on the Rule of Law Index estimations developed by the WJP. The well-regarded WJP Rule of Law Index is compiled from original surveys of the general public and local legal experts and measures nine dimensions of the rule of law, namely limited government powers, absence of corruption, order and security, fundamental rights, open government, effective regulatory enforcement, access to civil justice, effective criminal justice, and informal justice (12). This analysis used the component ‘effective regulatory enforcement,’ as the index item that most closely relates to the to mitigate the pandemic. The values of the index fall between 0 and 1, with 1 being the highest possible score (most effective regulation enforcement). In this analysis scores were transformed into point percentage to allow for meaningful interpretation of linear regression. Data are for the year 2020 (15).

Outpatient visits (per capita/year) was also included in the analyses to control for different health-care system utilization patterns. Data for the latest year available were retrieved from WHO dataset (16).

The analyses included countries of the WHO European region and USA and Canada as the only two North American members of Organization for Security and Cooperation in Europe (17).

We computed Pearson’s correlation coefficients including p-values between the pairs of variables. Multiple linear regression analysis was carried out to assess the independent association between explanatory variables and the outcome variable (excess death).

## Results

Mean excess mortality rate on the 2^nd^ of January 2022 among 50 countries included in the analysis was about 326±222 deaths per 100 000 population; mean vaccination rate was about 59%±18%; mean regulation enforcement was about 65±15 points percentage; and mean outpatients visits (per capita/year) was 6.3±2.5.

Excess death rate was strongly inversely correlated with regulation enforcement (r= −0.69 P<0.001) and vaccination coverage (r= −0.70, P<0.001). The later was strongly positively correlated with the regulatory implementation index (r=0.73, P<0.001).

No correlation was found between outpatient visits and each of three other variables in the analyses: excess mortality (r= 0.31 P=0.031), regulation enforcement (r=-0.13 P=0.406), or vaccination rate (0.001 P=0.997). The correlation of excess mortality and vaccination coverage for different scores of regulation enforcement is presented in Figure 1.

**Figure 1.**
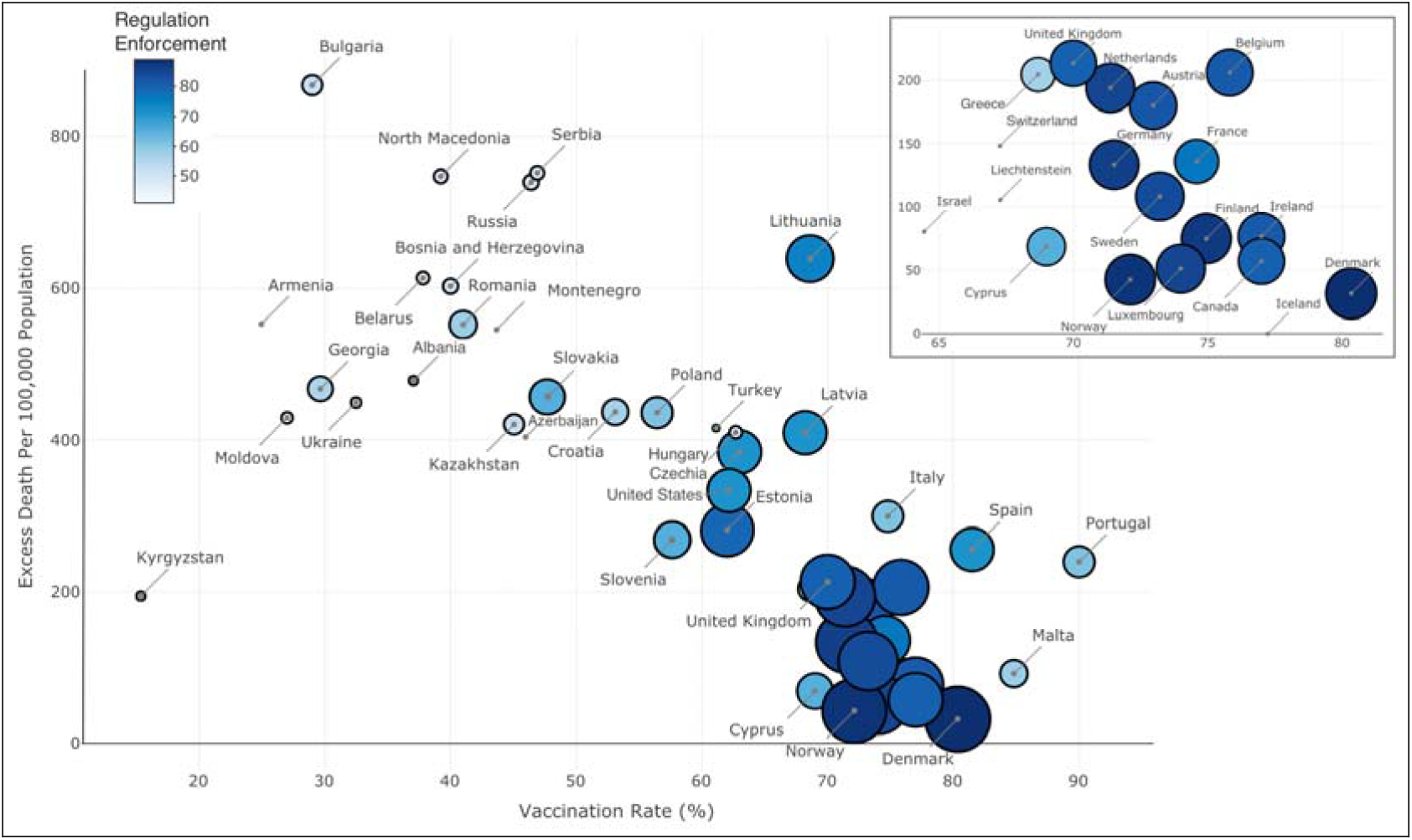
Association between COVID-19 related excess mortality, vaccination rate and regulation enforcement in 50 countries in January 2022. Regulation Enforcement with Excess Mortality −0.69 (p<0.001) Vaccination Rate with Excess Mortality −0.70 (p<0.001) Regulation Enforcement with Vaccination Rate 0.73 (p<0.001)

Excess mortality rate increased by 4.1/100000 for every percent decrease in vaccination rate, irrespective of regulation enforcement and health care utilization (Table 1). Furthermore, excess mortality increased with 6/100000 for every unit less in the regulatory implementation index score. Regulation enforcement, vaccination coverage, and outpatient visits explain about 62% of the variance of excess death (Table 1).

**Table 1.**
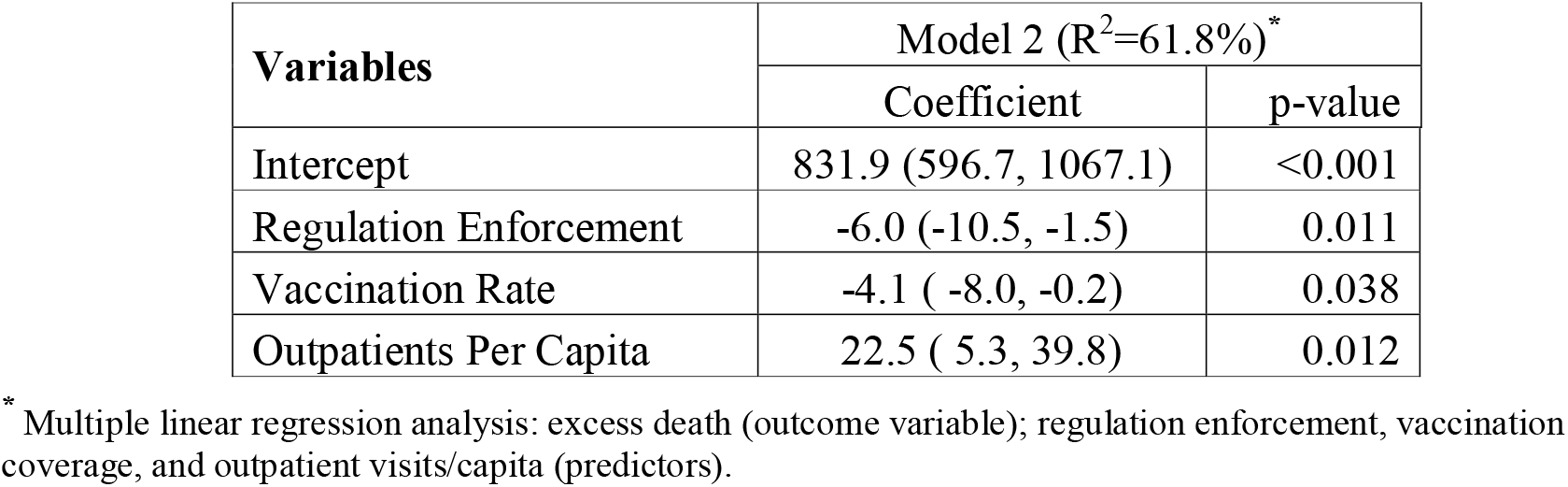
Association of excess mortality with regulation enforcement, vaccination coverage, and outpatient visits.

## Discussion

This short report provides a global perspective on comparative COVID-19 outcomes and policy solutions as of January 2022. In particular, the consideration of regulation enforcement differences provides a richer understanding of pandemic impact in the context of associated political and societal factors that can influence the management of the crises. Specifically, this study revealed that implementation of the not-always-popular government non-pharmaceutical interventions and, later, vaccination programs were more effective in more law abiding societies.

This provides key insights for Eastern Europe, which is understudied on this critical policy and public health topic. The focus of most international research and the global media has often focused primarily on outcomes of Western countries and associated factors – especially the resistance toward vaccination and public health measures (18) – pandemic outcomes have been poorer in Eastern Europe than Western counterparts. In the early phases of the pandemic, urgent short term measures were effective in preventing deaths in Eastern Europe (19), but over the longer term, other factors have played a more important role in mitigating the pandemic.

The degree of enforcement of government regulations is independently correlated to the excess mortality observed during 2020 and 2021 pandemic years. Regulation enforcement may have affected mortality directly, because of less effectively implemented social distancing regulations and mask mandates. This may be especially true in the pre-COVID vaccine period. In the second half of the 2020, during the second wave of COVID 19 in Europe when the vaccines were not yet a factor, excess mortality in many Eastern European countries begin to surpass the numbers recorded in the countries where pandemic first started (6). Regulation enforcement may also have affected mortality through a less successful vaccination program during 2021, as countries with a lower regulation implementation index have lower vaccination rates. Vaccine provision in EU countries has been centralized (20) and uniform for all members, while Russia was one of the first countries in the world to produce a vaccine (21). Hence, the lower success of vaccination campaigns in many Eastern European countries cannot be explained by variations in supply.

Regulation implementation may be influenced by several underlying social factors, including community trust, population education, public patience, etc. Other more specific factors related to present or past models of government may also play a role. Most Eastern European countries have been governed by authoritarian regimes in recent history and many citizens there may be more suspicious towards government regulations or mandates (22).

We discuss here only major factors related to regulation enforcement and implementation of vaccination programs. Other more complex factors may have interacted and a limitation of this analysis is the possibility of aggregation bias. Nevertheless, we expect this short report to encourage more discussion and especially further research in this policy area of regulation enforcement in this pandemic. We recommend that implementation enforcement factors should be taken into account by governments when considering population wide public health measures, and by experts when measuring real-world impact of the measures.

## Data Availability

All data produced in the present study are available online at Our World In Data database, WHO database and World Justice Project database

